# Estimating spatial disease rates using health statistics without geographic identifiers

**DOI:** 10.1101/2022.04.18.22274002

**Authors:** Javier Cortes-Ramirez, Juan D. Wilches-Vega, Ruby N. Michael, Vishal Singh, Olga M. Paris-Pineda

**Affiliations:** Centre for Data Science, Queensland University of Technology, Brisbane, Australia; Faculty of Medical Sciences and Health, University of Santander, Cúcuta, Colombia; Children’s Health and Environment Program, University of Queensland, Brisbane, Australia; Cities Research Institute, Griffith University, Brisbane, Australia; School of Public Health and Social Work, Queensland University of Technology, Brisbane, Australia

**Keywords:** Urban Sections, spatial epidemiology, Local Indicators of Spatial Autocorrelation -LISA, Empirical Bayes Smoothed Rate, Cucuta -North Santander, public health decision-making

## Abstract

Morbidity statistics can be reported as grouped data for health services rather than for individual residence area, especially in low-middle income countries. Although such reports can support some evidence-based decisions, these are of limited use if the geographical distribution of morbidity cannot be identified. This study estimates the spatial rate of Acute respiratory infections (ARI) in census districts in Cúcuta -Colombia, using an analysis of the spatial distribution of health services providers. The spatial scope (geographical area of influence) of each health service was established from their spatial distribution and the population covered. Three levels of spatial aggregation were established considering the spatial scope of primary, intermediate and tertiary health services providers. The ARI cases per census district were then calculated and mapped using the distribution of cases per health services provider and the proportion of population per district in each level respectively. Hotspots of risk were identified using the Local Moran’s I statistic. There were 98 health services providers that attended 8994, 18450 and 91025 ARI cases in spatial levels 1, 2 and 3, respectively. Higher spatial rates of ARI were found in districts in central south; northwest and northeast; and southwest Cúcuta with hotspots of risk found in central and central south and west and northwest Cucuta. The method used allowed overcoming the limitations of health data lacking area of residence information to implementing epidemiological analyses to identify at risk communities. This methodology can be used in socioeconomic contexts where geographic identifiers are not attached to health statistics.

## 1. Introduction

Effective decision making in public health is underpinned by using the best available evidence and research on risk factors determinants of disease. An important condition to conduct effective analysis in research is the availability and quality of data that can be used in various quantitative methodological frameworks (European Centre for Disease Prevention and Control, 2019). Statistics and health reports are especially important for epidemiological studies to identify the association of risk factors with health outcomes in the general population and support potential prevention strategies. However, statistical reports can be inadequate to provide the best evidence unless they can be used in meaningful analyses for at-risk populations (Kneale et al., 2018). Decisions related to diagnostic and therapeutic interventions are usually based on high-quality research studies such as clinical trials. In contrast, public health decisions, especially by local authorities, are often based on the review of basic statistics with limited academic rigour (McGill et al., 2015). The lack of high-quality data can favour the role that politics and political ideology have on shaping local governments initiatives in public health which can impact effective evidence-based decision making (Kneale et al., 2019). This can be accentuated in low and middle-income countries where the public health sector has limited access to resources for population health research (Kumar et al., 2018).

Public health departments can collect statistics from health providers such as hospitals, general practices and clinics and health insurance providers to monitor public health. The utility of these data depends on the inclusion of individual identifiers or categories such as socioeconomic status or residential areas. Public health data linked to small geographical areas such as counties or local governments are especially useful to identify the distribution of diseases or mortality in cities or larger regions (Cortes-Ramirez et al., 2020). In addition, census information for geographical areas can be linked to the public health data to calculate proportional rates of cases by population and the association of these proportions with other indicators such as socioeconomic and education indexes. This comes with challenges, in that basic health statistics may include unformatted and incomplete data and a lack of information at the individual level. This paucity of information is more evident in resource-constrained countries with up to 152 low-income and middle-income countries having no enough health data or poor-quality health data (Boerma & Stansfield, 2007).

The North Santander region in Colombia has an important burden of disease due to respiratory infections, with Cúcuta the capital city, accounting for more than 80% of the respiratory morbidity in the region (Gobierno de Colombia, 2021; Instituto Nacional de Salud, 2019). The city is located near the border with Venezuela (Figure 1) and its economy has been affected by the recent socioeconomic crises of this neighbouring country with important impacts on employment, security and poverty (Suárez González, 2016). Although the city is a commercial hub for the region’s mining activities and the production and exports from other economic activities (Ministerio de Comercio Turismo y Hoteleria, 2022), Cucuta had the second highest unemployment rate compared with other capital cities in 2019, and a 6.1% higher poverty level compared to the national average (Camara de Comercio de Cucuta, 2019; Departamento Adminstrativo de Planeacion, 2020). This has increased the pressure on the health sector considering the high demand of health services from people in condition of socioeconomic vulnerability and the increasing migrant population from Venezuela (Avendaño Sánchez, 2020).

**Figure 1.**
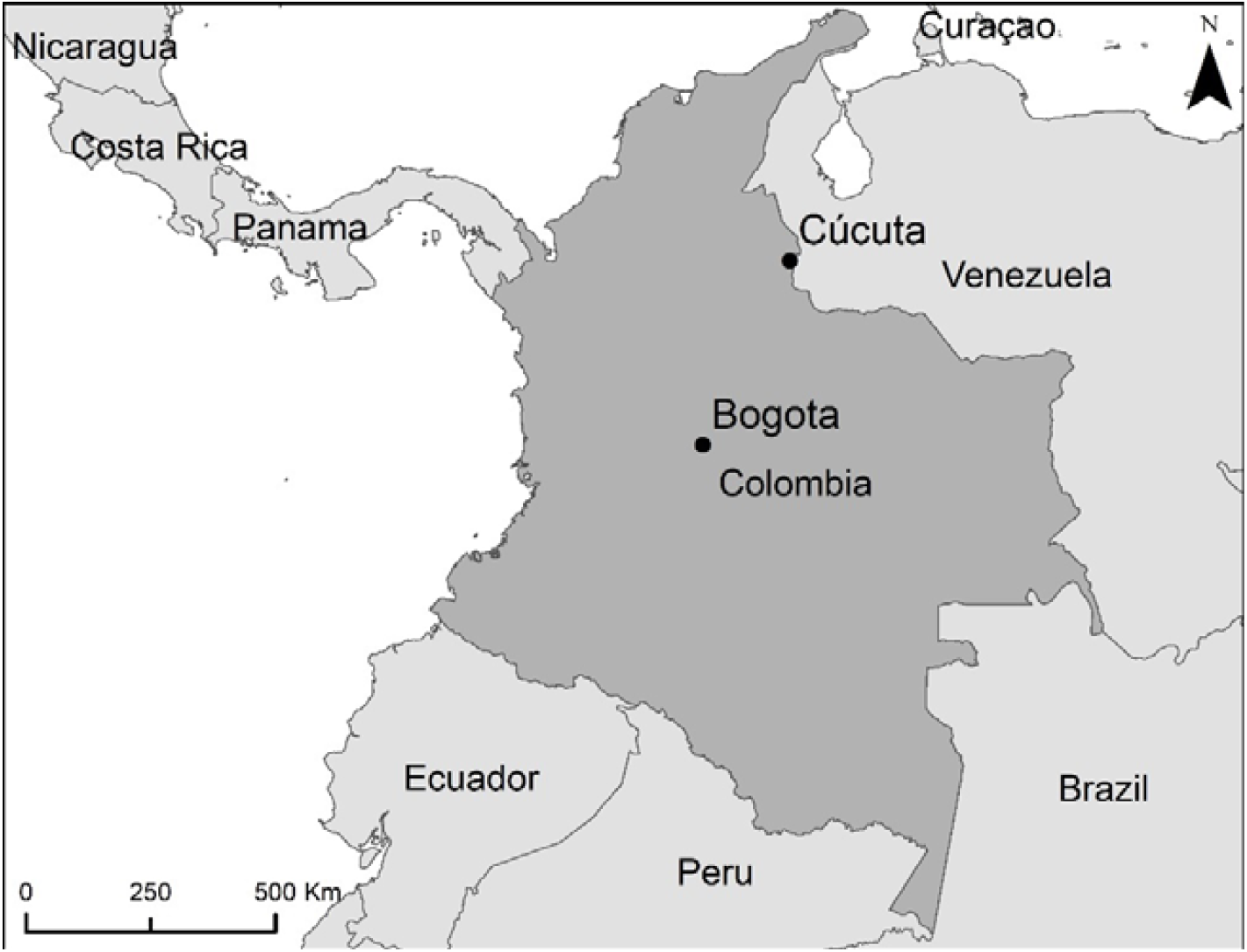
Geographical location of Cúcuta, Colombia.

The city has a complex network of public and private health services for primary, intermediate and tertiary healthcare. The population is distributed in geographical areas defined by census district boundaries called urban sections (USECs). Despite the organisation by USECs and the socioeconomic diversity between these districts, the statistics of acute respiratory infections (ARI) are often obtained only at the health centre and hospital level. The usability of statistics of ARI in Cúcuta for analyses to support evidence-based decision-making in public health is very limited because it is not linked to the distribution of the population in specific areas. The use of high-spatial-resolution health estimates including the neighbourhood and suburb levels has been demonstrated to be an effective approach to identify determinants of health issues than can be addressed from a public health perspective (Jain et al., 2021; Tao et al., 2022). However, the lack of geographic identifiers in the Cucuta’s ARI data prevents the calculation of health indicators with bigger impact for planning health strategies such as the rate of ARI per USEC.

Although the cases of ARI in Cúcuta are not reported at a geographical level, the geographical distribution of the health care providers (HCPs) can be used to make estimations of the number of cases occurring in each USEC. Spatial statistics can be used to estimate the distribution of observations in well-defined geographic settings using assumptions based on known characteristics of the population and health services to interpolate observations and compensate for the lack of better-quality data (Boakes et al., 2016; Griffith et al., 1989). A similar approach can be used to estimate the rate of ARI in the Cúcuta’s USECs, considering the spatial distribution of the HCPs and the population per USEC. The aims of this study are to design a method to calculate and map the spatial rate of ARI per USEC in Cúcuta, using statistics of cases reported at the HCP level, and to identify the hotspots of higher risk of ARI.

## 2. Materials and Methods

This study uses data on the number of medical consultations with a diagnosis of ARI in the period 01-January-2018 to 31-December-2018 categorised by HCP, obtained from the Cucuta’s Health Department. An initial analysis established a spatial scope (i.e., geographical area of influence) for each HCP and assigned them to three spatial level zones. Then, the ARI cases per USEC were calculated according to their location within these level zones and their proportional population per level zone. This study was approved by the ethics committee of the University of Santander, record VII-FT-025-UDES, 021 25/06/2019.

### 2.1 Study setting

The city of Cúcuta is located in the northeast of Colombia (Figure 1) and accounts for a population of 787,891 inhabitants of which 51.6% are female and 48.4% male (Alcaldia de San Jose De Cúcuta, 2021). Most of the population (68.8%) is economically active (16-64 year) with 22.8% population younger than 15 years and 8.4% older than 65 years. The city has an area of 1,176 km² with an average altitude of 320 mts above sea level. It is geographically organised in 460 USECs with an average size of 113,558 m2 (quartile-1: 83,216 m2; quartile-3: 175,929 m2) (figure 2) (Departamento Adminstrativo de Planeacion, 2021).

**Figure 2.**
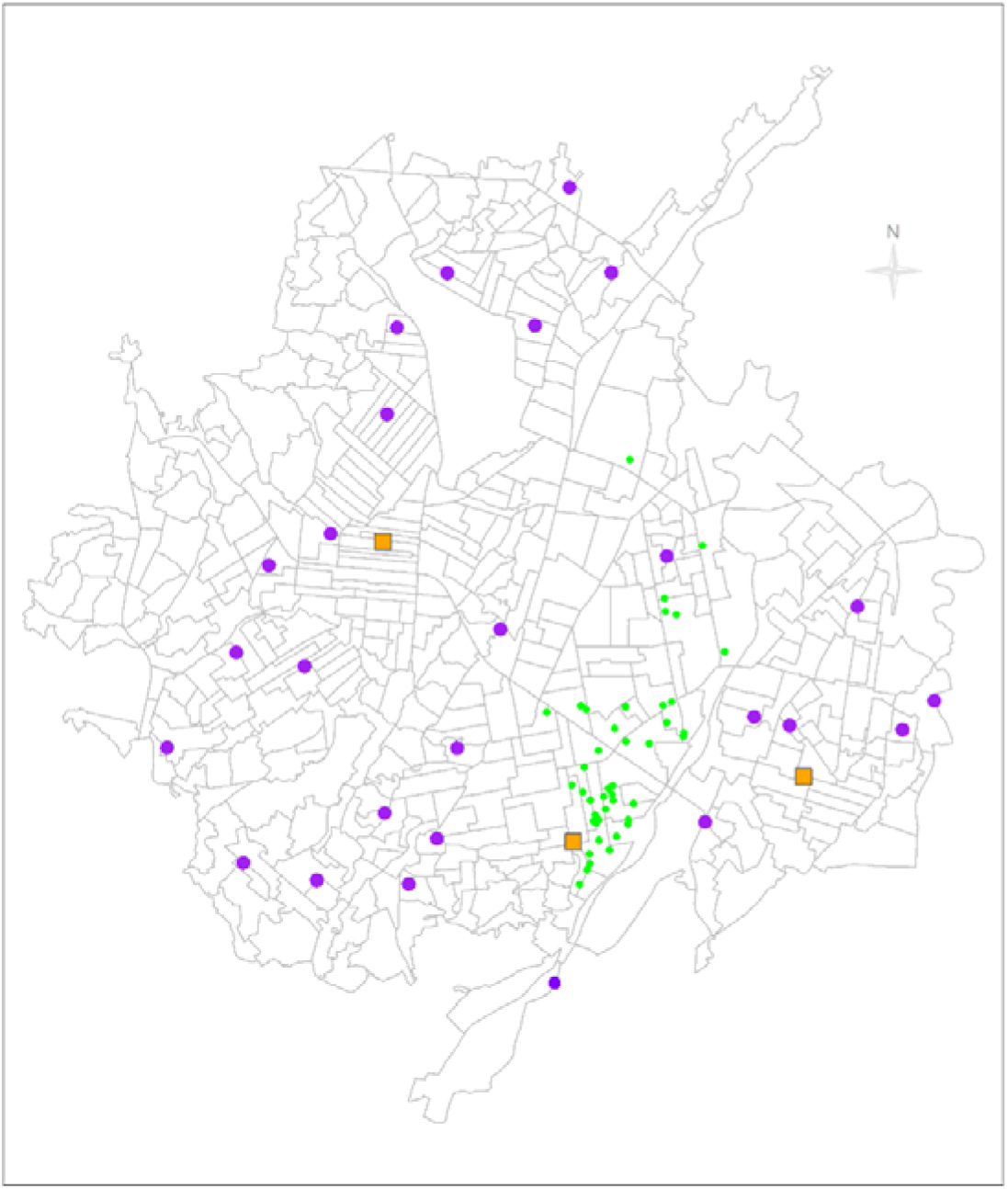
Spatial distribution of the Urban Sections in Cúcuta and the primary (purple dots) and intermediate (squares) public health care providers; and hospitals, private health services and GP clinics (green dots)

### 2.2 Spatial scope of the HCPs

The spatial scope of each HCP was established considering their spatial location and the complexity of their health services using the framework of the public health model: Primary HCPs provide basic health services to circumscribed areas and the population in their vicinity; intermediate HCPs provide more complex health services (including some procedures and medical specialities to populations in large areas of the city); and tertiary HCPs which are hospitals that provide high-complexity health services to the whole of the population (Congreso de la Republica, 2011). Whereas the boundaries of the vicinity of primary HCPs and larger areas serviced by intermediate HCPs are not established, the health system works on the assumption that the population access the closest health service (Tovar-Cuevas & Arrivillaga-Quintero, 2014). The private HCPs in contrast, are explicitly non-selective of the residence area, and although the majority are located in central areas (close to the commercial district), most of them provide services to the population in any area of the city. The private HCP categories were assigned using the same rationale: GP clinics, intermediate private services and private hospitals were included as primary, intermediate and tertiary HCPs respectively. Figure 2 shows the spatial distribution of the Cucuta’s USECs and HCPs.

### 2.3 Spatial zones

As there would be USEC within the spatial scope of more than one HCP, the spatial scope was assigned independently to spatial zones with different levels of spatial aggregation. Three level zones were established using the spatial scope of the public HCPs: Level 1 zones were the most tightly defined and included the USECs within the spatial scope of each public primary HCP. Level 2 zones included the USECs in the spatial scope of the public intermediate HCPs. The Level 3 zone included all Cúcuta’s USECs, defined as the spatial scope of the hospitals and the nonselective private HCPs (Figure 3). The definition of the spatial scope of HCPs in level 1 and 2 zones was based on the use of a nearest neighbour algorithm in ARCmap (v.10.6) (i.e., identifying the closest USECs to each HCP) and a preliminary analysis of the population served by each HCP. This included an assessment of the impact of the bus routes and the river shape to reflect accurately how the population access the HCPs services. Each level zone would have a total of ARI cases calculated as the count of cases of the HCPs within the zone. Since each HCP is assigned to a level zone only, there are not overlaps in the ARI cases count between HCPs.

**Figure 3.**
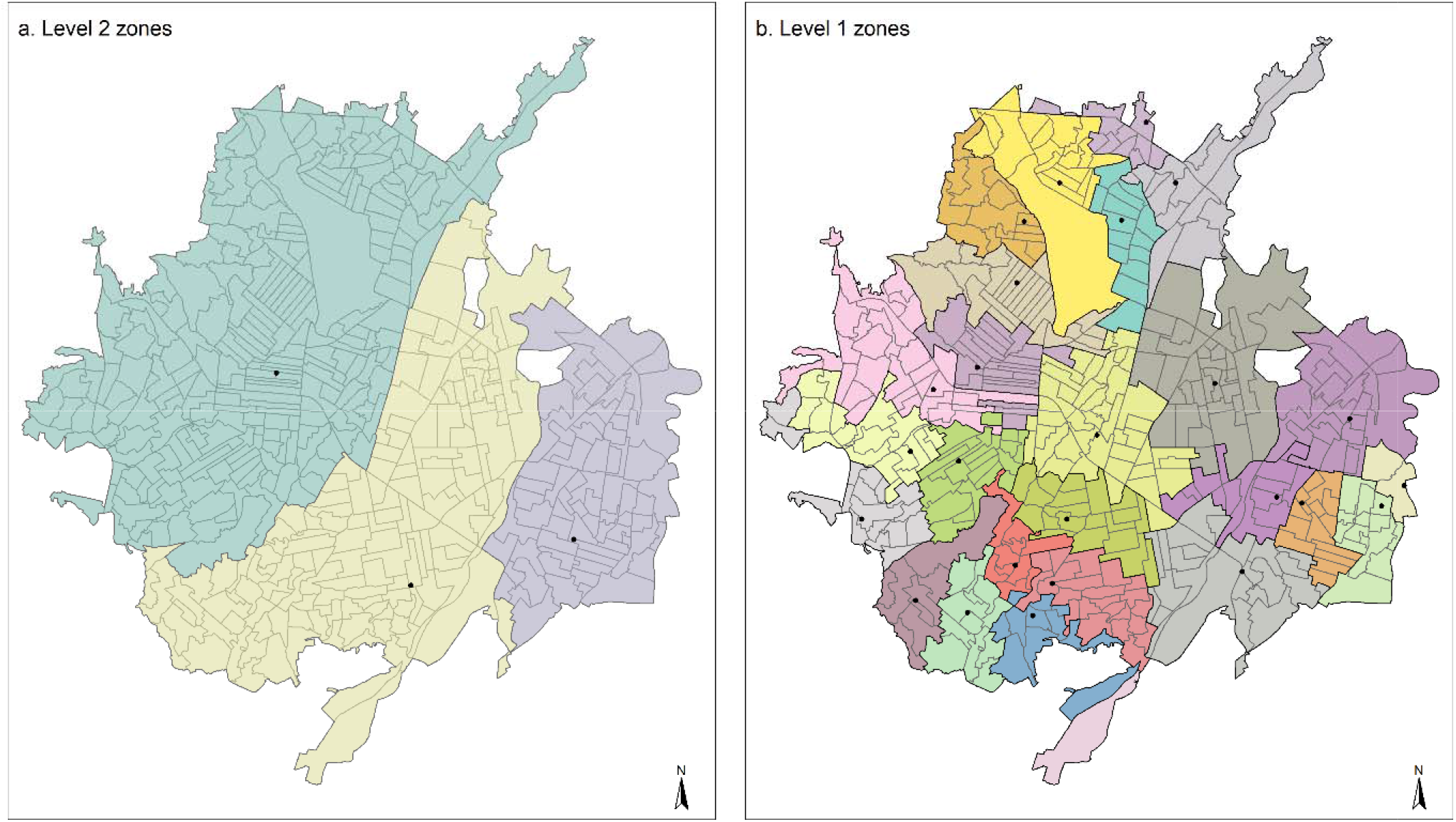
Spatial distribution of the geographic zones for levels of aggregation 2 (a) and 1 (b). The dots represent public intermediate and primary health care providers, respectively

### 2.4 Cases of ARI per USEC

To estimate the number of ARI cases per USEC, a weight value was assigned to each USEC according to the proportion of population in each level zone. For example, the weight value of a USEC with 500 people in a level 1 zone with a total population of 5000 people is 500 ÷ 5000 = 0.1. Each USEC would have ARI cases calculated separately in each of the three level zones, therefore, the final ARI cases per USEC were calculated as the sum of ARI cases in the three level zones.

### 2.5 Spatial smoothing

Once the total ARI cases were calculated for each USEC, the incidence rate of ARI per USEC could have been estimated as the total ARI cases divided by the USEC population. However, this rate would be calculated assuming an even spatial distribution of cases which can produce distortion of the estimates in addition to the lack of individual data to identify patients who could have attended more than one HCP. To overcome these issues, the ARI rate can be modelled using a spatial smoothing technique. Spatially smoothed rates borrow information from neighbour areas to increase the robustness of estimates by reducing variance instability potentially occurring in spatially organised data (Anselin et al., 2006). Spatial smoothing is conventionally implemented using Bayesian principles, particularly the Empirical Bayes (EB) Smoothed Rate approach (Anselin et al., 2010). The spatial rate of ARI per USEC (ARI-sr), were estimated in GeoDa (v. 1.18) using the spatial EB algorithm. The EB approach was also used to assess the spatial trend of ARI in Cúcuta to identify Local Indicators of Spatial Autocorrelation (LISA). These represent the USECs with significant higher ARI-sr surrounded by USECs also having a significantly higher ARI-sr (Anselin, 1995). Since the LISA identify USECs with higher ARI-sr compared to all USECs they can be interpreted as hotspots of relative risk of ARI. The local Moran’s I test in GeoDa was used to estimate the LISA of ARI in Cucuta. Maps were drawn in R using the R-package Tmap (Tennekes, 2018).

## 3. Results

There were 118,469 cases of ARI in Cúcuta over the study period, of which 37,060 (31%) were reported by primary health services or GP clinics; 53,677 (45%) were reported by intermediate HCP and 27,732 (23%) were reported by hospitals. There were 69 (70%) HCPs and GP clinics providing primary health services; 19 (19%) HCP providing intermediate health services and 5 (5%) hospitals (tertiary health services). Table 1 shows the number of ARI cases by HCP type and spatial zone level. All primary public HCPs had a spatial scope on spatial zones level 1. Ten (36%) of the primary private HCPs; 5 (33%) of the GP clinics and 5 (26%) of the intermediate HCPs (3 public and 2 private) had a spatial scope on spatial zones level 2. Fifteen (54%) of the primary private HCPs; 10 (66%) of the GP clinics; 15 (79%) of the intermediate HCPs and all hospitals had a spatial scope on spatial zone level 3.

**Table 1.** Cases of acute respiratory infections per Health Care Provider and spatial level of aggregation

| Health Care Provider (HCP) | N | cases | Spatial Zone |  |  |
| --- | --- | --- | --- | --- | --- |
|  |  |  | level 1 | level 2 | level 3 |
| Primary public HCP | 26 | 8994 | 8994 |  |  |
| Primary private HCP | 28 | 23726 |  | 2852 | 20874 |
| GP clinics | 15 | 4340 |  | 1179 | 3161 |
| Intermediate HCP | 19 | 53677 |  | 14419 | 39258 |
| Hospitals | 5 | 27732 |  |  | 27732 |
| Total | 98 | 118469 | 8994 | 18450 | 91025 |

Figure 3 shows the spatial zones for levels 1 and 2 of spatial aggregation. Three level 2 zones were identified from the spatial scope of the public intermediate HCPs (Figure 3a) and 26 level 1 zones were identified from the spatial scope of the public primary HCP (Figure 3b).

The count of cases of ARI in each spatial level zone are shown in Table 1. There were 8,994 (7.5%) ARI cases in level 1 zones (from public primary HCPs); 18,450 (15.6%) ARI cases in level 2 zones (from primary and intermediate HCP and GP clinics); and 91,025 (76,8%) ARI cases in the level 3 zone (from primary and intermediate HCPs, GP clinics and hospitals).

The distribution of the spatial rate of ARI (ARI-sr) is shown in Figure 4.a. The ARI-sr ranged from 0.16 to 3.65 per inhabitant, with higher ARI-sr found in USECs in the central south; northwest and northeast; and southwest regions. Three USECs in the central and north regions had more than one ARI case per inhabitant (i.e., ARI-sr >1).

**Figure 4.**
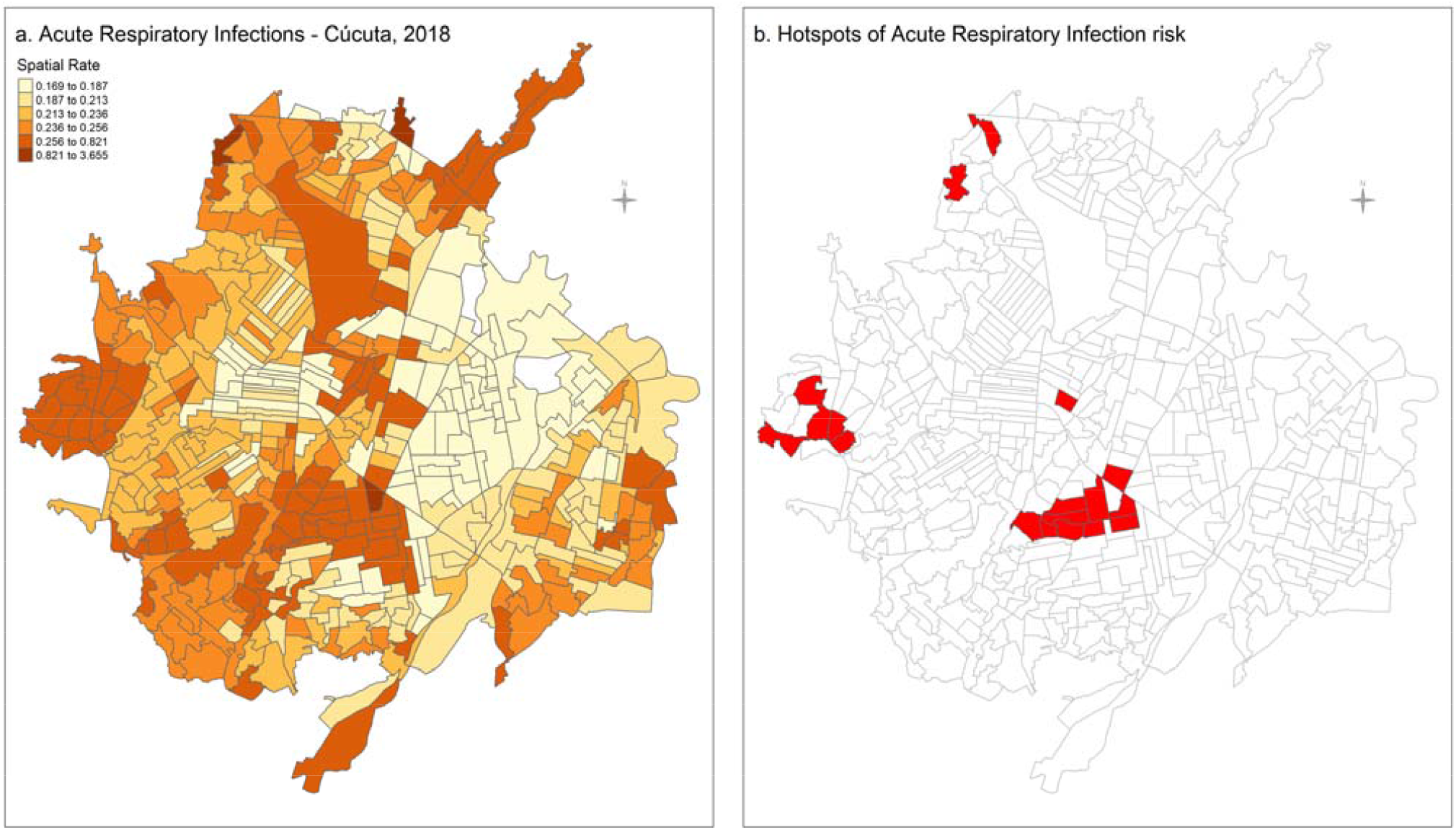
Spatial rates of Acute Respiratory Infections (a) and statistically significant hotspots of risk of Acute Respiratory Infections (b), in Cúcuta (2018).

The distribution of LISA of higher risk is shown in Figure 4.b. There were multiple hotspots of higher risk of ARI clustered together in central south and west Cúcuta and three isolated hotspots of higher risk in central and north Cúcuta.

## 4. Discussion

In this study, we calculated and mapped higher spatial rates of Acute Respiratory Infections (ARI-sr) in multiple urban sections (USEC) in central and north regions of Cúcuta and identified the hotspots of higher risk of ARI in the city. We first identified the USECs in the spatial area of influence of each health care provider (HCP) implementing a method to calculate the number of ARI cases in each USEC according to their population and the spatial scope of the HCP assigned to spatial levels of aggregation. The methodological approach allowed the estimation of morbidity rates in the Cucuta’s census districts despite the lack of geographical identifiers in the ARI data. This is the first study to estimate the rate of respiratory infections in small geographical areas in a major city in the Colombia northeast region.

This analysis identified the distribution of morbidity due to ARI with a considerable high spatial resolution as there were 460 spatial units (USECs) covering all geographical areas in Cucuta. While higher rates of ARI were found in USECs in all spatial regions of the city, the findings show a somewhat increased number of USECs with higher rates in the north, central and west regions. This was corroborated statistically with the identification of significant hotspots of risk in USECs in these regions. We modelled the spatial distribution of the ARI cases and the risk of ARI using assumptions on the population’s access to services of the health care providers according to their location and applying a Bayesian spatial analysis for risk estimation. Modelling data to overcome issues with imperfect vital data or missing health data for small areas has been used in disease mapping analyses in low- and middle-income countries in Latin America and Africa (Dwyer-Lindgren et al., 2018; Schmertmann & Gonzaga, 2018). The use of patterns of factors associated with the health data in combination with Bayesian spatial models has been increasingly used to fill gaps to allow a robust estimation of mortality indicators at a subnational level (Alexander et al., 2017; Local Burden of Disease H. I. V. Collaborators, 2021). These studies support important components of the method used in this analysis and provide a robust context for the incorporation of Bayesian statistics for disease mapping to support public health decision-making in regions with lack of high-quality health data.

The distribution of the risk in respiratory infections found in this study suggests that some factors can be more determinant in these areas. Having an indicator of morbidity on respiratory infections at the census districts level allows further assessment of their relationship with potential factors such as the index of socioeconomic development and some demographics characteristics including ethnicity, gender and age groups, for which data can be accessed from the census. This can increase the utility of the morbidity rates calculated in this study to produce more specific and complex analyses. For example, correlation tests can be used to explore predictive relationships between health outcomes and some risk factors of interest (Franco & Di Napoli, 2017) for better understanding the potential effect of environmental exposures on specific diseases and their relationship with public health interventions (Linka et al., 2020; Zartarian et al., 2017). The additional data on multiple factors can also be used for multivariate analysis to estimate the relative influence of one or more predictors on the ARI rates and to identify outliers or anomalies (Hu et al., 2019). This is particularly important in studies at a geographical level because of the potential risk of ecological bias, where confounding factors at the group level can produce spurious associations. The use of statistic techniques such as the design of mixed-effects spatial regressions using the spatial units as the random effects can reduce the risk of ecological bias (Cortes-Ramirez et al., 2022; Wakefield, 2009). This can be used in combination with the calculation of rates presented in this study and extended to identify potentially associated risk factors by designing further multivariate analyses.

The incorporation of a spatial dimension to calculate an indicator such as the risk of ARI allows a quick identification of specific communities more in need of public health interventions and can help better planning the implementation of strategies in the areas at high risk. The availability of relevant and adequate data on the morbidity due to respiratory infections is key for public health services to deal with many challenges including efficient health care provision and planning (Kneale et al., 2019). We used a GIS and geospatial software in combination with modelling of the characteristics of the Cúcuta’s health services and population’s distribution to estimate morbidity indicators in small areas. The use of these techniques and methodological approach is consistent with the increasing implementation of data analysis that considers the spatial distribution of health outcomes to provide robust measures to support decision making in the health sector (Alexander et al., 2017; Chinnaswamy et al., 2019; Local Burden of Disease H. I. V. Collaborators, 2021). The spatial rates of ARI at a suburban level estimated in this study would help targeting those communities with higher risk to prioritise the use of public health resources which can be particularly important in low- and middle-income countries. In addition, this methodology can support other analyses for strategic planning of suburban land use (Loibl & Toetzer, 2003) and understanding demographic trends in densely populated areas (Pan, 2021). Our findings show that the absence of health data classified at census districts or other administrative levels can be overcome using an analytical assessment of the spatial distribution of the health services and the population and using available software and statistics tools to support the implementation of meaningful epidemiological analyses. The potential distortion introduced by having assumptions about the distribution of cases in small urban areas can be reduced by the smoothing incorporated in spatial rate calculations to increase the robustness of the estimates (Blangiardo & Cameletti, 2015).

This analysis has some limitations, especially regarding the lack of data for specific individual markers such as age or gender that would have allowed the estimation of age and sex-adjusted spatial rates. The age-structure of the population can have great variability (Israëls, 2013) while some vulnerable groups can have a higher risk of IRA (Cortes-Ramirez et al., 2021). To reduce the potential effect of the age structure of the population, we calculated spatial rates which reduces the potential effect of outliers by smoothing the estimates across the USECs while taking into account the spatial trend or the crude rate (Anselin et al., 2006). Nevertheless, the standardisation of rates by age and or sex should be considered in further analyses using these data. In addition, we obtained health statistics of 2018 and matched these data with the 2018 census districts but did not incorporate a temporal analysis which would have allowed identifying a temporal trend, producing stronger estimates. However, this would be effective in contexts with a periodic census rather than the large gap of census data in Colombia. The previous census was in 2005 and the spatial trend of the census districts data is less reliable.

Another important limitation relates to the assumptions on the definition of USECs accessing health services of the HCPs. Whereas the use of 3 level zones decreases the impact of the assumptions not being met in all cases and spatial smoothing reduces the impact of outliers, the number of cases per USEC might not reflect an accurate count. As this is the main issue that this study aims to overcome it is important to consider that the rates and risk estimated are modelled approximations useful to support decision making and that future research using individual areas of residence is needed to provide further evidence on the outcomes of this study.

## 5. Conclusions

The limitations of health statistics that do not include area of residence to implementing epidemiological analyses to identify communities at higher risk can be overcome using spatial analyses that consider the spatial location of the health care providers. This study used a methodology to establish spatial levels of aggregation considering the geographical area of influence of health services to model the number of cases and estimate rates of acute respiratory infections in census districts in a capital city in Colombia, despite the absence of geographical identifiers in the health data. This method allowed the estimation of high-spatial resolution rates of these diseases which would increase the effectiveness of epidemiological studies to identify risk areas and support targeting resources for public health strategies and or interventions focused on higher risk communities. The methodological approach can be expanded to implement other relevant epidemiological analyses such as the identification of hotspots of risk and the implementation of multivariate analyses. The methodology developed in this study can be used to increase the usability and impact of public health statistics, especially in low- and middle-income countries where restrictions due to limited resources affect the availability of adequate and quality health data.

## Data Availability

All data produced in the study are available upon reasonable request to the authors. The raw data provided by the Department of Health is subject to their authorisation to be published.

## References

Alcaldia de San Jose De Cúcuta. (2021). Plan de Desarrollo 2020–2023. (Report 22062021). Departamento de Planeacion. https://cucuta.gov.co/pagina/wp-content/uploads/2021/10/51031_sgr-capitulo-alcaldia-municipal-de-san-jose-de-cucuta.pdf.

Alexander, M., Zagheni, E., & Barbieri, M. (2017). A Flexible Bayesian Model for Estimating Subnational Mortality. Demography, 54(6), 2025–2041. https://doi.org/10.1007/s13524-017-0618-7

Anselin, L. (1995). Local Indicators of Spatial Association—LISA. Geographical Analysis, 27(2), 93–115. https://doi.org/10.1111/j.1538-4632.1995.tb00338.x

Anselin, L., Lozano, N., & Koschinsky, J. (2006). Rate transformations and smoothing. Urbana, 51, 61801.

Anselin, L., Syabri, I., & Kho, Y. (2010). GeoDa: an introduction to spatial data analysis. In Handbook of applied spatial analysis (pp. 73–89). Springer.

Avendaño Sánchez, M. (2020). Cúcuta, política y economía frente a la migración venezolana Escuela de Economía, Administración y Negocios. Facultad de Negocios 舰].

Blangiardo, M., & Cameletti, M. (2015). Spatial and spatio-temporal Bayesian models with R-INLA. John Wiley & Sons.

Boakes, E. H., Fuller, R. A., McGowan, P. J. K., & Mace, G. M. (2016, 20s16/03/31). Uncertainty in identifying local extinctions: the distribution of missing data and its effects on biodiversity measures. Biology Letters, 12(3), 20150824. https://doi.org/10.1098/rsbl.2015.0824

Boerma, J. T., & Stansfield, S. K. (2007, 2007/03/03/). Health statistics now: are we making the right investments? The Lancet, 369(9563), 779–786. https://doi.org/10.1016/S0140-6736(07)60364-X

Camara de Comercio de Cucuta. (2019). Panorama economico regional. Informe del observartorio economico, Retrieved from http://datacucuta.com/images/panoramaeconomico2019.pdf.

Chinnaswamy, A., Papa, A., Dezi, L., & Mattiacci, A. (2019). Big data visualisation, geographic information systems and decision making in healthcare management. Management Decision.

Congreso de la Republica. (2011). Ley 1438 de 2011. Diario Oficial de Colombia, 47.957_19.01.2011.

Cortes-Ramirez, J., Vilcins, D., Jagals, P., & Soares Magalhaes, R. J. (2020). Environmental and sociodemographic risk factors associated with environmentally transmitted zoonoses hospitalisations in Queensland, Australia. One health (Amsterdam, Netherlands), 12, 100206–100206. https://doi.org/10.1016/j.onehlt.2020.100206

Cortes-Ramirez, J., Wilches-Vega, J. D., Paris-Pineda, O. M., Rod, J. E., Ayurzana, L., & Sly, P. D. (2021, Apr). Environmental risk factors associated with respiratory diseases in children with socioeconomic disadvantage. Heliyon, 7(4), e06820. https://doi.org/10.1016/j.heliyon.2021.e06820

Cortes-Ramirez, J., Wraith, D., Sly, P. D., & Jagals, P. (2022, Jan 21). Mapping the Morbidity Risk Associated with Coal Mining in Queensland, Australia. Int J Environ Res Public Health, 19(3). https://doi.org/10.3390/ijerph19031206

Departamento Adminstrativo de Planeacion. (2020). Plan de Desarrollo Municipal. Alcaldia de San Jose de Cucuta, Retrieved from http://ieu.unal.edu.co/images/Planes_de_Desarrollo_2020/C%C3%BAcuta_37396_3--pdm-san-jose-de-cucuta-2020--2023-v31-07052020.pdf.

Departamento Adminstrativo de Planeacion. (2021). Sistema Estadístico Nacional. (Reporte: La información del DANE en la toma de decisiones regionales. Cúcuta, Norte de Santander). https://www.dane.gov.co/files/investigaciones/planes-departamentos-ciudades/210319-InfoDane-Cucuta-Norte-de-Santander.pdf

Dwyer-Lindgren, L., Squires, E. R., Teeple, S., Ikilezi, G., Allen Roberts, D., Colombara, D. V., Allen, S. K., Kamande, S. M., Graetz, N., Flaxman, A. D., El Bcheraoui, C., Asbjornsdottir, K., Asiimwe, G., Augusto, Â., Augusto, O., Chilundo, B., De Schacht, C., Gimbel, S., Kamya, C., Namugaya, F., Masiye, F., Mauieia, C., Miangotar, Y., Mimche, H., Sabonete, A., Sarma, H., Sherr, K., Simuyemba, M., Sinyangwe, A. C., Uddin, J., Wagenaar, B. H., & Lim, S. S. (2018, 2018/08/13). Small area estimation of under-5 mortality in Bangladesh, Cameroon, Chad, Mozambique, Uganda, and Zambia using spatially misaligned data. Population Health Metrics, 16(1), 13. https://doi.org/10.1186/s12963-018-0171-7

European Centre for Disease Prevention and Control. (2019). The use of evidence in decision-making during public health emergencies. Stockholm: ECDC. https://doi.org/DOI:10.2900/63594

Franco, F., & Di Napoli, A. (2017, 2017/04/01). Measures of Association in Medicine and Epidemiology. Giornale di Tecniche Nefrologiche e Dialitiche, 29(2), 127–128. https://doi.org/10.5301/GTND.2017.16951

Gobierno de Colombia. (2021). Morbilidad consulta externa. Medicina General. 2019. Sistema de Datos Abiertos, Retrieved from https://www.datos.gov.co/browse?Informaci%C3%B3n-de-la-Entidad_Departamento=Norte+de+Santander&Informaci%C3%B3n-de-la-Entidad_Municipio=C%C3%BAcuta&category=Salud+y+Protecci%C3%B3n+Social&page=2.

Griffith, D. A., Bennett, R. J., & Haining, R. P. (1989, 1989/11/01). Statistical Analysis of Spatial Data in the Presence of Missing Observations: A Methodological Guide and an Application to Urban Census Data. Environment and Planning A: Economy and Space, 21(11), 1511–1523. https://doi.org/10.1068/a211511

Hu, Y., Yu, S., Qi, X., Zheng, W., Wang, Q., & Yao, H. (2019). An overview of multiple linear regression model and its application. Zhonghua yu Fang yi xue za zhi [Chinese Journal of Preventive Medicine], 53(6), 653–656.

Instituto Nacional de Salud. (2019). Infección respiratoria aguda. Semana epidemiológica 40. Boletin Epidemiologico Semanal, Retrieved from: https://www.ins.gov.co/buscador-eventos/BoletinEpidemiologico/2019_Boletin_epidemiologico_semana_40.pdf.

Israëls, A. (2013). Methods of standardisation. The Hague/Heerlen, The Netherlands: Statistics Netherlands.

Jain, S., McLoughlin, C., Cooney, J., McLoughlin, A., Abdalla, A., & MacHale, S. (2021). Suburban vs urban: do the attendee’s demographic profile influence the emergency department’s mental health characteristics presentation? BJPsych Open, 7(S1), S327–S328. https://doi.org/10.1192/bjo.2021.861

Kneale, D., Rojas-García, A., & Thomas, J. (2018). Exploring the importance of evidence in local health and wellbeing strategies. Journal of Public Health, 40(suppl_1), i13–i23. https://doi.org/10.1093/pubmed/fdx152

Kneale, D., Rojas-García, A., & Thomas, J. (2019, 2019/06/28). Obstacles and opportunities to using research evidence in local public health decision-making in England. Health Research Policy and Systems, 17(1), 61. https://doi.org/10.1186/s12961-019-0446-x

Kumar, M., Gotz, D., Nutley, T., & Smith, J. B. (2018). Research gaps in routine health information system design barriers to data quality and use in low- and middle-income countries: A literature review [Review]. The International journal of health planning and management, 33(1), e1–e9. https://doi.org/10.1002/hpm.2447

Linka, K., Peirlinck, M., & Kuhl, E. (2020, 2020/10/01). The reproduction number of COVID-19 and its correlation with public health interventions. Computational Mechanics, 66(4), 1035–1050. https://doi.org/10.1007/s00466-020-01880-8

Local Burden of Disease H. I. V. Collaborators. (2021, Jan 8). Mapping subnational HIV mortality in six Latin American countries with incomplete vital registration systems [Article]. BMC Med, 19(1), 4. https://doi.org/10.1186/s12916-020-01876-4

Loibl, W., & Toetzer, T. (2003, 2003/07/01/). Modeling growth and densification processes in suburban regions—simulation of landscape transition with spatial agents. Environmental Modelling & Software, 18(6), 553–563. https://doi.org/10.1016/S1364-8152(03)00030-6

McGill, E., Egan, M., Petticrew, M., Mountford, L., Milton, S., Whitehead, M., & Lock, K. (2015). Trading quality for relevance: non-health decision-makers’ use of evidence on the social determinants of health. BMJ Open, 5(4), e007053. https://doi.org/10.1136/bmjopen-2014-007053

Ministerio de Comercio Turismo y Hoteleria. (2022). Perfil economico y comercial del departamento Norte de Santander. Estudios Economicos, Retrieved from https://www.mincit.gov.co/estudios-economicos/perfiles-economicos-por-departamentos.

Pan, T. (2021). A Health Support Model for Suburban Hills Citizens. Applied System Innovation, 4(1). https://mdpi-res.com/d_attachment/asi/asi-04-00008/article_deploy/asi-04-00008.pdf?version=1611907166

Schmertmann, C. P., & Gonzaga, M. R. (2018). Bayesian Estimation of Age-Specific Mortality and Life Expectancy for Small Areas With Defective Vital Records. Demography, 55(4), 1363–1388. https://doi.org/10.1007/s13524-018-0695-2

Suárez González, E. J. (2016). Diagnóstico de la situación del desarrollo económico de Cúcuta durante las dos últimas administraciones, como base para la creación de una Zona de Régimen Aduanero Especial en la ciudad Universidad del Rosario].

Tao, Y., Ma, J., Shen, Y., & Chai, Y. (2022, 2022/10/01/). Neighborhood effects on health: A multilevel analysis of neighborhood environment, physical activity and public health in suburban Shanghai. Cities, 129, 103847. https://doi.org/10.1016/j.cities.2022.103847

Tennekes, M. (2018). tmap: Thematic Maps in R. Journal of Statistical Software, 84(1), 1–39.

Tovar-Cuevas, L. M., & Arrivillaga-Quintero, M. (2014). Estado del arte de la investigación en acceso a los servicios de salud en Colombia, 2000-2013: revisión sistemática crítica. Revista Gerencia y Políticas de Salud, 13(27), 12–26.

Wakefield, J. (2009, Apr). Multi-level modelling, the ecologic fallacy, and hybrid study designs. Int J Epidemiol, 38(2), 330–336. https://doi.org/10.1093/ije/dyp179

Zartarian, V., Xue, J., Tornero-Velez, R., & Brown, J. (2017, Sep 12). Children’s Lead Exposure: A Multimedia Modeling Analysis to Guide Public Health Decision-Making. Environ Health Perspect, 125(9), 097009. https://doi.org/10.1289/EHP1605

